# Enhancing molecular and genomic approaches for *Bordetella pertussis* investigation and public health surveillance in Victoria, Australia

**DOI:** 10.64898/2026.10.11.26364704

**Authors:** Lamali Sadeesh Kumar, Michelle Sait, Muhamad Fachrul, Louise M. Judd, Jake A. Lacey, Janet Strachan, Sarah L. Baines, Benjamin P. Howden, Norelle L. Sherry

## Abstract

*Bordetella pertussis* is the main aetiological agent of pertussis (whooping cough), with a global resurgence of cases reported in 2024. As current pertussis diagnostics are predominantly based on molecular methods, cultured isolates are increasingly scarce, limiting traditional culture-based surveillance. To address these challenges, this study evaluated culture-independent approaches for *B. pertussis* surveillance and their feasibility for implementation in public health laboratory settings in Victoria, Australia. The Microbiological Diagnostic Unit Public Health Laboratory received 356 nasopharyngeal clinical specimens that were PCR-positive for *Bordetella* spp. from diagnostic laboratories for further characterisation during the 2024-2025 epidemic period. Culture-based approaches yielded only five *B. pertussis* isolates, largely due to limitations associated with specimen transport media. Four culture-independent approaches were evaluated for *B. pertussis* characterisation: real-time PCR assay for high-resolution *Bordetella* spp. differentiation (*n*=356), multiplex PCR amplicon sequencing assay for lineage typing (*n*=38), in-house laboratory developed test implemented on Panther Fusion® System for macrolide resistance screening (*n*=92) and shotgun metagenomic sequencing (*n*=5). The dataset was dominated by *B. pertussis* (330/356; 92.7%) with most strains harbouring *ptxP3* (35/38; 92.1%) allele and the macrolide resistance rate was reported as 4.3% (4/92). Shotgun metagenomics was able to detect *B. pertussis* directly from clinical samples, but the genomic recovery was limited. Evaluation of operational performance and workflow integration highlighted method-specific strengths and limitations, informing suitability for implementation in public health laboratories. This study demonstrates the potential to integrate culture-independent approaches into public health laboratory settings to strengthen *B. pertussis* surveillance and inform feasible workflow design amid limited culture availability.

**Importance statement:** Culture-independent approaches are increasingly valuable for *Bordetella pertussis* investigations and surveillance, particularly as declining culture recovery limits the availability of isolates for downstream characterisation. The emergence and dissemination of vaccine-evading and macrolide-resistant *B. pertussis* strains highlight the need for robust laboratory strategies to support continuous surveillance and enhance preparedness for future pertussis epidemics. This study evaluates four culture- independent molecular and genomic approaches for *B. pertussis*, including species detection and differentiation, macrolide resistance screening and lineage typing while assessing their operational characteristics and feasibility for implementation in public health laboratory settings. These findings provide insights into the potential role of integrated workflows in strengthening *B. pertussis* surveillance and public health investigations.

**Data summary:** Genome sequences of *Bordetella pertussis* isolates are deposited in GenBank under BioProject PRJNA870170. Sample data and accession numbers are included in the Supplementary Table S1. The authors confirm all supporting data, code and protocols have been provided within the article or through supplementary data files.

## Introduction

Pertussis (also known as ‘whooping cough’) is a highly contagious respiratory disease caused by *Bordetella pertussis*. Despite widespread vaccination programs and high immunisation coverage, many countries continue to experience pertussis epidemics every three to five years, indicating the ongoing circulation of *B. pertussis* worldwide. A significant resurgence of pertussis was reported in 2024, with multiple countries, including Australia, experiencing epidemics after a marked reduction in pertussis cases during the COVID-19 pandemic [1–3].

Pertussis initially presents with non-specific prodromal symptoms resembling a common cold, and can progress to paroxysmal coughing episodes, a hallmark clinical feature of the disease. Infants are particularly vulnerable and may develop serious complications of pertussis, including apnoea, cyanosis and death [4]. Laboratory diagnosis of pertussis is performed on nasopharyngeal specimens using *B. pertussis* culture and/or nucleic acid amplification testing (NAAT), typically PCR-based assays. However, diagnostic test accuracy heavily depends on disease stage at sampling, with culture most sensitive within the first 2 weeks of cough onset, while NAATs have highest sensitivity within the first 3 weeks and may remain positive for up to 5 weeks or longer [5, 6].

Given their higher sensitivity and extended detection window compared with culture, PCR-based approaches have largely replaced culture as the primary laboratory diagnostic method for pertussis. Multiplex PCR assays have further improved diagnostic workflows by enabling simultaneous detection of *Bordetella* spp. and other respiratory pathogens directly from clinical specimens [6–8]. The multicopy insertion sequence IS*481* is a widely used molecular target for *B. pertussis* detection, including in commercial assays, due to its high copy number (>200 copies per genome), which enhances analytical sensitivity. However, IS*481* is not entirely specific to *B. pertussis*, highlighting the need for complementary targets and appropriate assay design to ensure reliable differentiation from other *Bordetella* spp. Other *Bordetella* species, including *B. parapertussis*, *B. holmesii* and *B. bronchiseptica* (less commonly) can also cause pertussis-like respiratory illness in humans. Accurate differentiation of these species from *B. pertussis* is essential for reliable assessment of pertussis disease burden and vaccine impact, as these pathogens differ in clinical significance and their potential response to vaccine-induced protection [4, 6, 9, 10].

Ongoing surveillance of circulating *B. pertussis* lineages is essential for identifying changes in population structure, detecting emerging strains, assessing vaccine effectiveness and guiding evidence-based public health interventions. Despite widespread vaccination, pertussis continues to re-emerge globally, driven by factors including waning vaccine-induced immunity, changes in vaccine acceptance and emergence and dissemination of *B. pertussis* lineages with modified virulence factors that may evade vaccine-induced immunity [2, 6, 11–13]. Variation in the pertussis toxin promoter (*ptxP*), has driven global shifts in circulating *B. pertussis* populations, with ancestral *ptxP1*-harbouring lineages being progressively replaced by *ptxP3*-harbouring lineages associated with potentially increased pertussis toxin production [2, 6, 11]. *B. pertussis* exhibits very low genomic diversity, making lineage discrimination challenging for many typing schemes. Hence, multiple typing schemes have been developed and used globally for lineage typing, with particular emphasis on profiling acellular pertussis vaccine antigen genes and epidemiological markers to monitor the emergence and spread of vaccine-adapted lineages [2, 14–18]. However, the widespread adoption of PCR-based diagnostics has reduced the availability of *B. pertussis* isolates, limiting culture-based phenotypic and genotypic characterisation. Therefore, recent studies have adopted culture-independent approaches such as amplicon sequencing, targeted sequencing and shotgun metagenomics to investigate population structure and monitor circulating lineages [2, 12, 19].

Pertussis is treated with macrolides, primarily erythromycin or azithromycin and macrolide-resistant *B. pertussis* (MRBP) was first identified in the United States of America in 1994 [20]. Macrolide resistance is reported to be associated with an A→G substitution in the 23S rRNA gene at position 2037 relative to the *B. pertussis* Tohama I genome (accession no. NC_002929.2), equivalent to position 2047 in a previous version of the *B. pertussis* Tohama I reference genome [21, 22]. MRBP has been reported in various regions in recent years [11, 13, 23, 24], with marked resistance rates reported from China after the COVID-19 pandemic [13, 24]. Prevalence of MRBP remains low in Australia, with the first case reported in 2024 [25] and a resistant rate of 4% during the 2024 pertussis epidemic [2]. MRBP was predominantly associated with the *ptxP*1 lineage in earlier reports but has shifted to predominance in the *ptxP*3 lineage after the COVID-19 pandemic [2, 11–13, 22, 24, 25]. However, limited *B. pertussis* culture availability for phenotypic antimicrobial susceptibility testing and the lack of routine MRBP detection assays hinder timely surveillance of macrolide resistance and emerging resistant lineages.

With declining *B. pertussis* isolate availability, the adoption of culture-independent approaches is increasingly important for supporting public health genomic and molecular surveillance. However, appropriate method selection is critical to balance reliable performance, scalability and implementation feasibility within public health laboratory settings. Here, we present an overview of culture-independent approaches for investigating *B. pertussis*, with emphasis on accurate detection and differentiation of *Bordetella* spp., characterisation of circulating lineages and detection of macrolide resistance. We further evaluate operational performance and workflow integration of those approaches to support effective molecular and genomic surveillance of *B. pertussis* in public health laboratory settings.

## Materials and Methods

### Study design and clinical specimens

The study was performed at the Microbiological Diagnostic Unit Public Health Laboratory (MDU PHL) in the state of Victoria, Australia (population 7.1 million at 31^st^ December 2025 [26]). Clinical specimens testing positive for *Bordetella* spp. by PCR at diagnostic microbiology laboratories in Victoria were referred to MDU PHL for inclusion in this study. Clinical specimens collected between April and November 2024 and March 2025 were included in the study. Duplicate clinical specimens were identified and excluded, with the earliest specimen available for each patient included in the study.

### Culture-based testing

#### B. pertussis culture

All clinical specimens included in this study were cultured for *B. pertussis* on charcoal agar supplemented with cephalexin and incubated under aerobic conditions at 37 °C, with plates examined daily for up to 7 days. Recovered isolates were identified using VITEK® MS PRIME MALDI-TOF and only isolates identified as *B. pertussis* were included in the study.

#### Whole genome sequencing and bioinformatic analysis

DNA extraction from *B. pertussis* isolates was performed using the Virus/Pathogen DSP mini kit on the QIAsymphony (QIAGEN). DNA libraries were prepared using the Nextera XT kit following manufacturer’s instructions (Illumina). Whole genome sequencing was performed on the Illumina NextSeq 500/550 systems using 2 x 150 bp paired-end chemistry following manufacturer’s instructions.

Bohra bioinformatics pipeline (v3.7.2) (https://github.com/MDU-PHL/bohra) was used for quality control of sequence reads and genome assembly. SKESA (v2.5.1) (https://github.com/ncbi/SKESA) was used as the *de novo* sequence assembler, run with default parameters [27]. For lineage typing, the genomic targets subsequently interrogated by the amplicon sequencing assay were identified and aligned from genome assemblies using gene-puller (minimum 95% nucleotide identity and 90% coverage) against reference target sequences from *Bordetella pertussis* Tohama I (NC_002929.2). Resulting alignments were visualised in Geneious Prime (v2024.0.7) (http://www.geneious.com) to identify lineage-defining single nucleotide polymorphisms (SNPs) as previously described [12]. Assemblies were also screened for the A2037G mutation in 23S rRNA, associated with macrolide resistance in *B. pertussis* using AMRFinderPlus (v4.2.7) [28] with *B. pertussis* as the species option.

### Culture-independent testing

#### DNA extraction from clinical specimens

DNA extraction was performed for all clinical specimens included in this study using the QIAsymphony DSP Virus/Pathogen Midi Kit on the QIAsymphony platform (QIAGEN), incorporating a spike-in internal qPCR extraction control (Meridian Bioscience). Extracted DNA was used for downstream *Bordetella* spp. differentiation and *B. pertussis* lineage typing methods.

#### Real-time PCR assay for *Bordetella* spp. detection and differentiation

All clinical specimens included in this study were tested using a multiplex real-time PCR assay for detection and differentiation of *B. pertussis*, *B. parapertussis*, and *B. holmesii*, as previously described [10]. The assay comprised two multiplex PCR reactions targeting *IS481*/*IS1001* and *recA*/*IS1002*, respectively. Detailed assay conditions and *Bordetella* spp. differentiation criteria are provided in the Supplementary Methods (Supplementary Method 1). All reactions included qPCR Extraction Control RED – Control Mix 25× (Meridian Bioscience) as an internal extraction and amplification control.

#### Multiplex PCR amplicon sequencing and bioinformatic analysis for lineage typing

A multiplex PCR amplicon sequencing method was used for lineage typing following a previously described scheme targeting major *B. pertussis* lineage-associated alleles (*ptxP*, *fim3* and *)aB3*) and markers associated with Australian epidemic lineages (EL1–EL5) [12]. Detailed assay conditions are provided in the Supplementary Methods (Supplementary Method 2). Following confirmation by the *Bordetella* spp. differentiation assay described above, 38 *B. pertussis* positive clinical specimens spanning the collection period were included to evaluate the lineage typing approach.

Raw sequencing reads were assessed for quality using FastQC and summarised with MultiQC [29]. Adapter trimming was performed using Cutadapt. Trimmed paired-end reads were mapped to reference amplicon sequences using BWA-MEM. Resulting BAM files were sorted and indexed using SAMtools [30]. Lineage-associated SNPs were interrogated directly from mapped reads using SAMtools mpileup [30], and allele frequencies were calculated from nucleotide counts at predefined genomic positions. Alleles were assigned when supported by ≥10 reads and ≥70% of reads at the interrogated position. Mixed allele profiles were defined when a secondary allele was detected at ≥10% frequency and supported by ≥10 reads. Mapped reads were visually inspected in Geneious Prime (v2024.0.7) where required.

This lineage typing scheme first classified strains by the *ptxP* allele (*ptxP3* or non-*ptxP3*), followed by the *fim3* allele (*fim3A* or *fim3B*) within each *ptxP* group. Strains with the *ptxP3*–*fim3A* allelic profile were further assigned to Australian epidemic lineages (ELs) 1–5 [12]. This scheme also included detection of the *)aB3* allele, which was used as a proxy marker for macrolide-resistant *B. pertussis* strains belonging to the *ptxP1* lineage in the original assay design. [12].

#### Panther Fusion® laboratory developed test for macrolide resistance screening

A real-time PCR laboratory-developed test (LDT) was implemented on the Panther Fusion® System (Hologic) using the Open Access functionality for detection of the macrolide resistance-associated A2037G mutation in the *B. pertussis* 23S rRNA gene [31]. Following confirmation by the *Bordetella* spp. differentiation assay described above, a subset of 92 *B. pertussis* positive clinical specimens spanning the collection period were included to evaluate the macrolide resistance screening assay. This assay enabled direct testing of clinical specimens with DNA extraction, amplification and detection integrated on the same platform.

#### Shotgun metagenomic sequencing and bioinformatic analysis

Shotgun metagenomic sequencing was conducted on all clinical specimens that were confirmed positive for *B. pertussis* by culture at the Centre for Pathogen Genomics at the University of Melbourne (Doherty Institute). DNA extraction was performed using the EZ2 Powerfecal Pro DNA/RNA Kit (QIAGEN), and libraries were created using the Illumina DNA Prep Kit with one-quarter of the recommended reagent volumes. Libraries were sequenced on Illumina Nextseq 1000/2000 platform. ZymoBIOMICS Spike-in Control II (Low Microbial Load) (Zymo Research), diluted 1:10, was used as the internal control.

Raw paired-end reads from the sequencer were quality filtered and adapter trimmed using fastp (v1.0.1) [32], with quality assessed using FastQC and MultiQC [29]. Trimmed reads were mapped to the ZymoBIOMICS Spike-in Control II reference using Bowtie2 (v2.5.4) [33] and spike-in abundance and coverage metrics were calculated using SAMtools [30].

Human reads were removed using Hostile (v2.0.2) [34] followed by NoHuman (v0.5.0) [35]. Host-depleted reads were taxonomically classified using Kraken2 (v2.1.6; confidence threshold 0.1) against the PlusPF and GTDB reference databases. Species-level abundance estimates were generated using Bracken (v3.0.1) [36]. To quantify *B. pertussis,* reads assigned to taxid 520 were used to calculate reads per million (RPM).

Host-depleted reads were also mapped to *B. pertussis* Tohama I reference genome (NC_002929.2) using Bowtie2 (--very-sensitive –no-discordant –no-mixed –no-unal), reads mapping to multiple loci (MAPQ < 20) were excluded, and genome coverage statistics were calculated using SAMtools. Reads mapping to the *B. pertussis* reference genome were subsequently extracted and aligned to the target loci included in the *B. pertussis* lineage typing scheme using Bowtie2 (--very-sensitive --all). Marker- level metrics, including read count, depth, and breadth of coverage, were calculated using SAMtools.

Metagenomic assembly was performed on the reads mapping to *B. pertussis* independently per sample using metaSPAdes (SPAdes v4.0.0, ‘--metà mode) [37] with default k-mer length. Assembly contigs ≥1 kb in length were retained and binning was performed independently with MetaBat2 (v2.18) [38] and MaxBin2 (v2.2.7) [39], both using the re-mapped coverage depth. Bin sets from both binners were integrated and dereplicated using DAS Tool (v1.1.7) [40] with default score thresholds to produce a non-redundant set of refined MAGs. MAG completeness, contamination, and strain heterogeneity were assessed with CheckM2 (v1.1.0) [41] using the uniref100.KO.1.dmnd database, and chimeric contigs were identified using GUNC (v1.0.6) [42] against the ProGenomes2 reference database. Taxonomic assignment of MAGs was performed with GTDB-Tk (v2.6.1) [43] using GTDB release r226. Per-MAG quality metrics from all three tools were merged into a unified QC table.

### Operational assessment and workflow integration of culture-independent approaches

Operational assessment was conducted based on estimated cost per sample, turnaround time, and overall operational complexity. Overall operational complexity of each approach was assessed qualitatively based on the operational input variables including sample preparation requirements, instrument dependency, bioinformatics/data analysis requirements and hands-on labour time. Hands-on labour time was estimated based on cumulative operator time required from sample receipt to result reporting, excluding instrument run time and other passive processing periods. Estimated costs were based on reagents and consumables requirements excluding labour costs and turnaround time (TAT) was defined as the interval from sample receipt to result reporting. Estimated per-sample costs and TATs were quantified for each individual approach and for the integrated workflow where applicable.

Workflow integration was evaluated by assessing the role of each culture-independent approach in supporting public health surveillance of *B. pertussis*. Integration was further assessed by considering the placement of each approach within the testing pathway (standalone or reflex testing) and the scope of testing (targeted or untargeted).

## Results

### Clinical specimen characteristics and recovery of *B. pertussis* isolates

The study included a total of 356 residual clinical specimens that tested PCR-positive for *Bordetella* spp. at six diagnostic microbiology laboratories in Victoria. The specimens comprised 355 nasopharyngeal swabs and one sputum sample (Table S1).

Culture recovery yielded five viable *B. pertussis* isolates (1.4%; 5/356), including four from specimens transported in Liquid Amies Transport Medium and one from a specimen transported in Universal Transport Medium (UTM) (Table S1).

### *Bordetella* spp. distribution

*Bordetella* species differentiation by real-time PCR identified *B. pertussis* as the predominant species, accounting for 330 (92.7%) of the 356 specimens. Three specimens were positive for *B. holmesii* (0.8%), two of which were also positive for *B. pertussis*. *B. parapertussis* was not detected. Detection of the IS*481* target alone was consistent with the presence of *Bordetella* spp. (undifferentiated) in 17 (4.8%) specimens and no PCR targets were detected in 8 (2.2%) specimens. All clinical specimens with an IS*481* Ct value ≤30 also tested positive for the secondary target IS*1002*, confirming the presence of *B. pertussis* (Table S1). The spike-in internal control (Meridian Bioscience) was detected in all reactions, confirming successful DNA extraction and PCR amplification.

### Lineage diversity of *B. pertussis*

Multiplex PCR amplicon sequencing of 38 *B. pertussis*-confirmed specimens revealed that 35/38 (92.1%) strains carried the *ptxP*3 allele, while the remaining three were non-*ptxP*3. Among the strains harbouring *ptxP3* (*n*=35), 26 carried the *fim3B* allele, eight carried the *fim3A* allele and one specimen exhibited both *fim3A* and *fim3B* alleles. All three non-*ptxP*3 strains carried the *fim3*A allele (Figure 1; Table S1). None of the strains carried *)aB*3 allele (Table S1).

**Figure 1.**
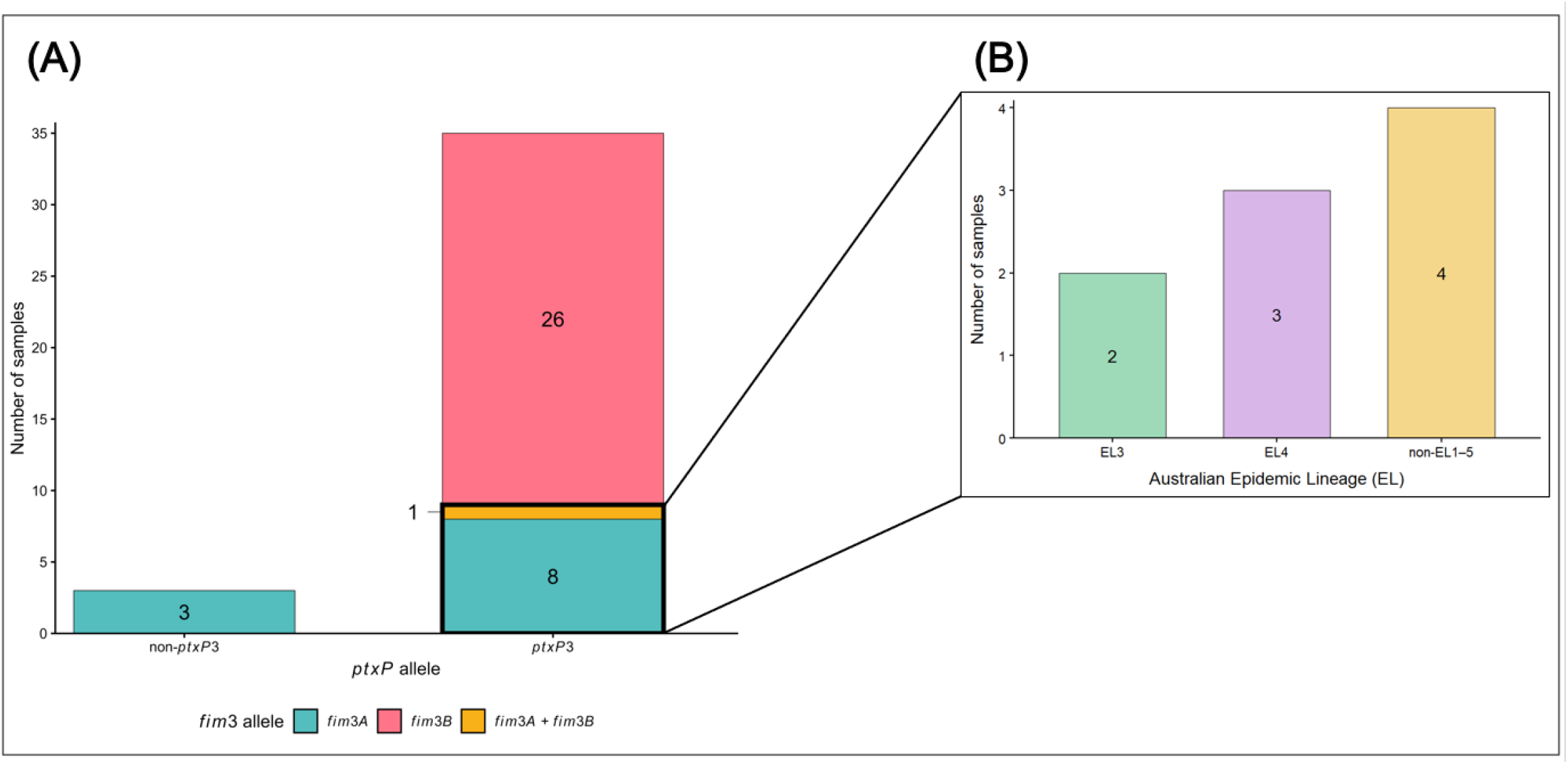
*ptxP* and *fim3* allelic profiles and lineage assignment among 38 *Bordetella pertussis*- positive specimens. (A) Distribution of *ptxP* and *fim3* alleles identified by multiplex PCR amplicon sequencing among 38 *B. pertussis*-confirmed specimens. The mixed profile indicates one specimen containing both *fim3A* and *fim3B* alleles. (B) Expanded view of the *ptxP3–fim3A* allelic profile (*n*=9, including the specimen with mixed *fim3* profile), showing Australian epidemic lineage (EL) assignment according to the previously described typing scheme [12]: EL3 (*n*=2), EL4 (*n*=3), and non-EL1–5 (*n*=4).

Among strains with the *ptxP*3–*fim3*A allelic profile (*n* =9), including one specimen with a mixed *fim3* profile, two Australian epidemic lineages (ELs): EL3 (*n* =2) and EL4 (*n* =3) were reported. The remaining four strains were not assigned to any Australian epidemic lineages defined in the scheme and were classified as non-EL1–5 (Figure 1; Table S1).

Whole-genome analysis of *B. pertussis* isolates (*n* =5) identified four isolates with the *ptxP3*–*fim3B* allelic profile and one isolate with the *ptxP3*–*fim3A* allelic profile, which was assigned to EL3 (Table S1).

### Emergence of macrolide resistance *B. pertussis* (MRBP) in Victoria

The A2037G mutation in 23S rRNA associated with macrolide resistance was detected in four of 92 (4.3%) *B. pertussis*-positive clinical specimens collected in 2024 by the Panther Fusion® LDT. The four MRBP strains corresponded to those exhibiting the *ptxP3*–*fim3A* allelic profile and classified as non- EL1–5 by the lineage typing scheme. The *)aB*3 allele was not detected in any MRBP strains (Table S1).

Whole-genome analysis of *B. pertussis* isolates (*n* = 5) confirmed that none harboured the A2037G mutation in the 23S rRNA gene (Table S1).

### Shotgun metagenomic sequencing results

All five clinical specimens that yielded *B. pertussis* by culture underwent shotgun metagenomic sequencing. Sequencing generated ∼18–33 million raw reads per sample, which were reduced following quality filtering and two rounds of host depletion, resulting in a wide range of total reads (∼27,000–22.4 million reads). Kraken2 detected *B. pertussis* (NCBI taxid 520) in all specimens and Bracken analysis showed *B. pertussis* relative abundances ranging from 0.00001-0.23 across samples (Table S2).

When mapped against the *B. pertussis* Tohama I reference genome (NC_002929.2) by bowtie2, mean read depth ranged from 0.03-5.57× while genome breadth of coverage ranged from 1.06%–91.24% across samples. AUSMDU00116112 specimen yielded the highest mean read depth (5.57x) and genome breadth of coverage (91.24%) (Table S3). A single metagenome assembled genome (MAG) was recoverable in AUSMDU00116112, which was classified as *B. pertussis,* however, it only had 34.67% completeness (Table S4). When *B. pertussis* reads were mapped against the targets used in the amplicon sequencing assay, mean read depths remained below 10× for all targets across all specimens (Table S3). Therefore, reliable lineage typing could not be performed, despite high breadth of coverage for some targets.

### Operational assessment and workflow integration of culture-independent approaches

The culture-independent approaches evaluated in this study served distinct roles, differing in information generated, operational requirements and suitability for integration within public health laboratory workflows. Shotgun metagenomic sequencing served as a standalone approach, while the other three approaches provided complementary information for species detection and differentiation, macrolide resistance screening and lineage typing of *B. pertussis* within an integrated workflow.

Operational assessment of the four culture-independent methods demonstrated that shotgun metagenomic sequencing had the highest estimated cost of A$350 per sample and a turnaround time of 2-3 days (Figure 2). In contrast, the integrated workflow combining the remaining three approaches had a substantially lower estimated cost of A$63 per sample, with shared DNA extraction reduced costs relative to the cumulative cost of individual approaches (Figure 2) and a similar turnaround time of 2-3 days. Shotgun metagenomic sequencing was the most operationally demanding approach, requiring specialised sequencing infrastructure, extensive bioinformatic processing and the highest hands-on labour time (∼5–6 hours). Despite incorporating multiple approaches, the integrated workflow demonstrated lower operational complexity and reduced hands-on labour time (∼4.5 hours) compared with shotgun metagenomic sequencing.

**Figure 2:**
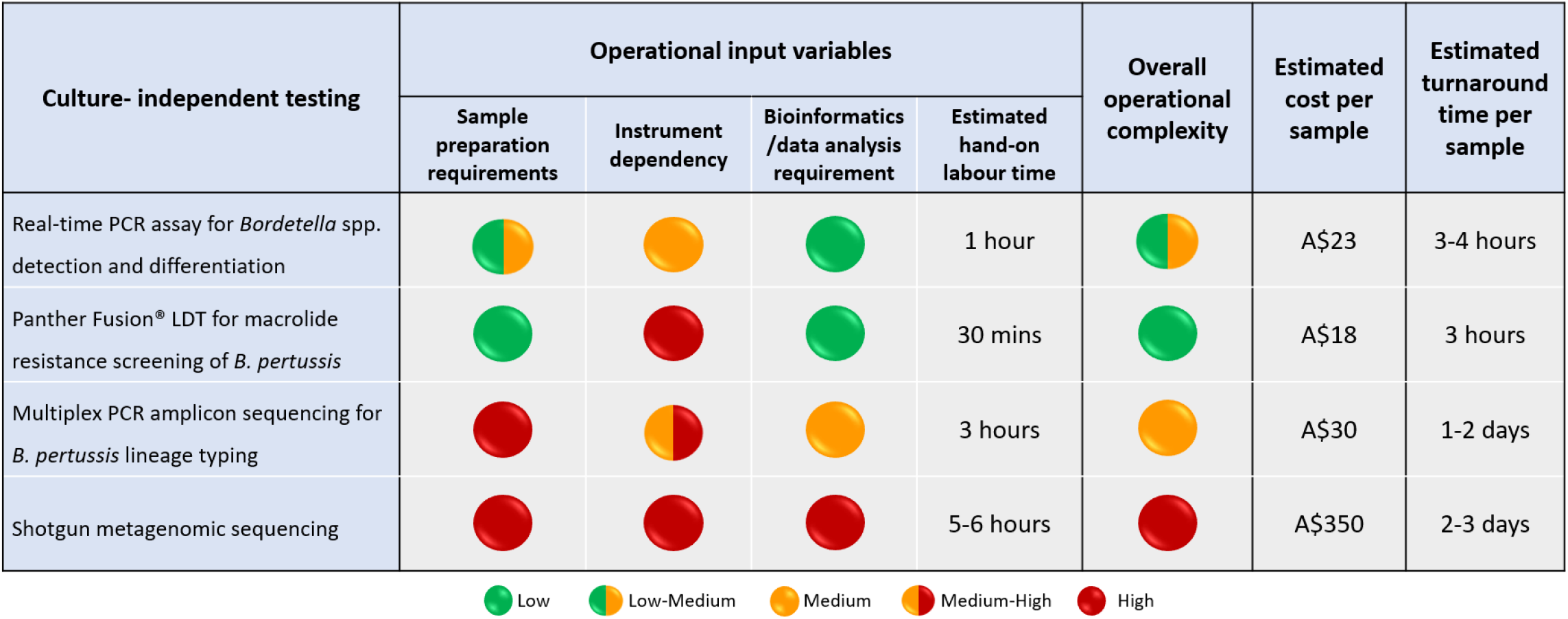
Operational assessment of culture-independent testing used in this study. Comparison of culture-independent testing for *Bordetella pertussis* based on laboratory operational burden, estimated cost per sample and estimated turnaround time per sample. Each method was evaluated independently as a complete end-to-end workflow from sample receipt to result reporting, including all pre-analytical and analytical steps (eg: DNA extraction, library preparation, sequencing and bioinformatics analysis where applicable). Overall operational complexity was assessed qualitatively based on sample preparation requirements, instrument dependency and bioinformatics/data analysis requirements, together with quantified hands-on labour time.

Evaluation of workflow integration identified a sequential testing pathway incorporating species detection and differentiation (standalone testing), with *B. pertussis*-positive specimens undergoing reflex testing for antimicrobial resistance and lineage typing within a single workflow suitable for public health laboratory settings. In contrast, shotgun metagenomic sequencing remained an exploratory standalone approach, requiring further optimisation before implementation in public health laboratory settings (Figure 3).

**Figure 3:**
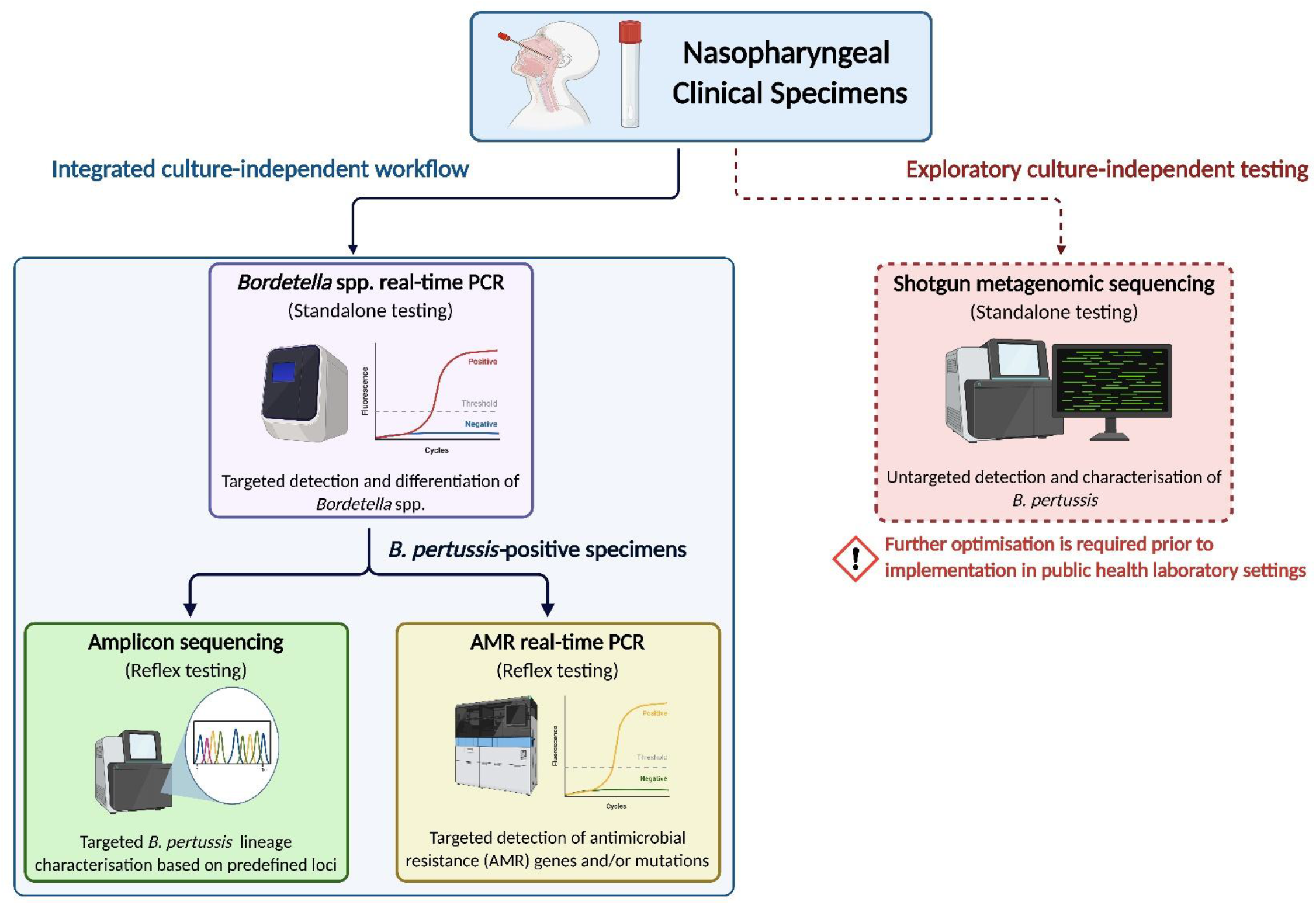
Conceptual framework for culture-independent workflow for *Bordetella pertussis* surveillance in public health laboratories. This evaluation-informed framework outlines a workflow for integrating culture-independent approaches to support *Bordetella pertussis* surveillance in public health laboratories from nasopharyngeal clinical specimens. The proposed workflow includes real- time PCR-based *Bordetella* spp. differentiation, with *B. pertussis*-positive specimens proceeding to reflex molecular screening for targeted antimicrobial resistance (AMR) markers and lineage typing by amplicon sequencing. Lineage typing would ideally align with a standardised *B. pertussis* typing framework to support consistent genomic surveillance. Shotgun metagenomic sequencing represents an exploratory approach for broader genomic characterisation requiring additional optimisation prior to routine application. Solid lines indicate the proposed integrated workflow, while dashed lines indicate exploratory pathways.

## Discussion

The growing adoption of culture-independent testing for *B. pertussis* diagnostics and surveillance has created a need to evaluate their practical implementation in routine public health laboratories for accurate species identification, detection and monitoring of macrolide resistance, and characterisation of *B. pertussis* population structure [2, 9, 10, 12, 19]. In this study, we applied four culture-independent testing approaches for the investigation of *B. pertussis* using direct clinical specimens collected during the most recent pertussis outbreak in Victoria, Australia, which began in 2024. We also assessed the feasibility of implementing these approaches in a public health laboratory setting.

Accurate identification of *B. pertussis* from other *Bordetella* spp. and characterising population structure is important in public health for assessing vaccine effectiveness and monitoring antimicrobial resistance [9, 10]. Most clinical specimens included in this study had been initially tested using commercial respiratory panels in diagnostic laboratories. However, given possible limitations in species-level resolution, a four-target real-time PCR assay [10] was applied to differentiate *B. pertussis* from *B. parapertussis* and *B. holmesii*. The predominance of *B. pertussis* (92.7%) observed in this dataset was anticipated, given that the specimens were collected during a pertussis epidemic period and was consistent with observations reported elsewhere [2, 3]. This assay detected *B. holmesii*, including a co-infection with *B. pertussis*, which may be missed by commercial respiratory panels. Such co-infections have been reported globally and highlight the importance of ongoing surveillance of non-pertussis *Bordetella* spp. associated with pertussis-like illness [12, 23, 44, 45]. Nevertheless, although macrolides are commonly used empirically for *B. pertussis* infections, previous studies have reported reduced macrolide susceptibility in *B. holmesii* and potential induced erythromycin resistance in *B. holmesii* and *B. parapertussis*, underscoring the importance of accurate identification of *Bordetella* spp. for appropriate antimicrobial treatments [46–48]. No *Bordetella* spp. was detected in eight specimens submitted as PCR-positive for *Bordetella* spp., potentially reflecting low bacterial load, specimen degradation or differences in assay sensitivity. While this study was conducted during a period of heightened *B. pertussis* activity, further evaluation of *Bordetella* spp. circulation during non-epidemic periods would provide additional insights into these findings.

Understanding circulating lineages in the population, particularly during pertussis epidemics, is important for monitoring changes in *B. pertussis* population structure and supporting public health surveillance. Although based on a relatively small subset of samples, *ptxP3* allele was predominant (92.1%; 35/38) in this dataset, suggesting ongoing expansion of vaccine-adapted and more virulent strains in Victoria, in line with trends observed across Australia in recent years [2, 12, 15]. However, *ptxP3-fim3B* (or *ptxP3*-*fim3*-2 [15]) allelic profile was prevalent in the 2024 pertussis epidemic in this dataset and other Australian studies [2] while *ptxP*3-*fim3*A (or *ptxP3-fim3-*1 [15]) allelic profile was prevalent in the two previous epidemics in Australia since 2007, prior to the COVID-19 pandemic [12, 15]. Recent phylogenetic studies have highlighted that most post-pandemic strains reported in Australia were related to pre-pandemic circulating lineages, except for macrolide-resistant strains, suggesting expansion of pre-existing lineages rather than emergence of novel ones [11]. Other *B. pertussis* lineage typing studies have incorporated additional vaccine antigen genes and epidemiological markers for strain characterisation [2, 11, 24], however, the targeted nature of Australian lineage typing scheme used in this study limited the number of shared markers available for direct comparison across studies. While multiple schemes exist for *B. pertussis* lineage typing [14, 16–18, 49], adoption of a globally recognised typing scheme would improve comparability across studies and support coordinated international surveillance efforts.

Studies conducted after the COVID-19 pandemic have reported increased emergence and dissemination of macrolide-resistant *B. pertussis* (MRBP) carrying the A2037G mutation in the 23S rRNA gene in Australia and multiple countries across Asia, Europe, and North America. These recent MRBP strains often associated with the *ptxP3*–*fim3*A (or *ptxP3-fim3-*1 [15]) allelic profile [2, 11, 13, 23–25], which was also observed among the MRBP strains (*n*=4) identified in this study. However, the MRBP strains in this study were not assigned to Australian epidemic lineages (ELs) 1–5 assessed by the lineage typing scheme [12], and further evaluation of their relatedness was limited by the lack of fine-scale phylogenetic resolution. WGS was not possible due to the lack of isolates and shotgun metagenomic sequencing was not applied due to anticipated limited genomic recovery. Although the*)aB3* allele has previously been described as a proxy marker for MRBP strains belonging to the *ptxP1* lineage in China [12, 22], its utility as a marker was not evident in this study, where all MRBP strains belonged to the *ptxP3* lineage. While macrolide resistance rate remains low in this study (4.3%; 4/92) and other Australian reports [2], recent genomic surveillance from China has reported a marked increase in resistance among *B. pertussis* isolates, rising from less than 50% before the COVID-19 pandemic to nearly 100% in the post-pandemic period [11, 13, 24]. These findings highlight the need to integrate routine MRBP testing into ongoing surveillance in Australia.

Shotgun metagenomic sequencing approach used in this study detected *B. pertussis* in all five specimens, however, the data were insufficient for metagenome assembled genome (MAG) reconstruction or lineage typing. The variability in *B. pertussis* read counts and relative abundance across the five specimens highlights challenges associated with metagenomic detection in specimens with low and uneven pathogen loads [50]. Given the low genetic diversity of *B. pertussis*, adequate sequencing depth is crucial to ensure reliable variant calling and lineage resolution. Deep shotgun metagenomic sequencing and targeted culture-independent sequencing approaches have shown improved *B. pertussis* genomic recovery from clinical specimens in recent studies [2, 12, 19]. Moreover, sample handling including transport, storage, and host depletion strategies, requires careful consideration to improve downstream genomic recovery [51, 52].

When evaluating culture-independent testing approaches for *B. pertussis* in public health laboratory settings, considerations such as cost, turnaround time, workflow integration, and resource requirements are some of important factors influencing implementation. In this study, it is important to note that operational estimates are context dependent and may vary according to sample number per batch, instrument throughput, and laboratory workflow configuration, particularly for sequencing- based methods where cost and turnaround times are strongly influenced by run utilisation. The approaches evaluated in this study provided distinct intended applications, with some functioning as standalone tests and others better suited for integration into diagnostic workflows. A combined workflow integrating *B. pertussis* detection, macrolide resistance screening and lineage typing enables detailed characterisation of *B. pertussis* strains from clinical specimens. These approaches are cost-effective, rapid and have low-medium operational complexity, making those more readily adaptable to public health laboratories in Australia. Furthermore, automated platforms such as the Panther Fusion® System may facilitate implementation in public health laboratories by reducing hands-on labour requirements and leveraging existing automated molecular testing infrastructure. However, as targeted assays, these methods may require ongoing refinement to ensure detection of emerging resistance mechanisms and newly circulating lineages within surveillance systems. While untargeted metagenomic and targeted sequencing approaches are increasingly applied in diagnostic testing and have the potential to provide comprehensive analysis of *B. pertussis* strains [2, 12, 19], their high cost and high operational complexity remain key limitations for routine implementation. The application of these approaches may also differ depending on the surveillance objective, with untargeted metagenomics suited for broad pathogen detection and targeted approaches being more practical for species-specific strain characterisation. Continued improvements in culture-independent sequencing technologies, supported by capacity building and workflow optimisation are likely to enhance their feasibility for implementation in public health laboratory settings [50, 53].

Evaluation of *B. pertussis* culture recovery in this study highlighted the challenges associated with recovering viable organisms from clinical specimens collected under current diagnostic workflows. Most specimens were transported in media that may have contained antimicrobial agents and were designed for molecular detection of a wide range of respiratory pathogens. Combined with delayed culture due to the retrospective study design, these factors may have significantly reduced *B. pertussis* viability and culture recovery [6, 25]. However, strategies to improve culture recovery, such as dedicated culture swabs and optimised transport conditions could be challenging to implement across a statewide network involving multiple pathology providers.

In conclusion, this study demonstrates the importance of working towards integration of culture- independent approaches into public health laboratory settings to strengthen *Bordetella pertussis* surveillance, thus mitigating limited culture availability. Observing the continued circulation of vaccine adapted strains and the emergence of macrolide-resistant *B. pertussis* in this study underscores the need to review current diagnostic and surveillance strategies to inform public health responses and preparedness for future pertussis epidemics. This study provides practical guidance for the implementation of culture-independent testing approaches and workflows design for *B. pertussis* investigations in public health and reference laboratory settings.

## Data Availability

All data produced in the present work are contained in the manuscript, and genome data available online at NCBI SRA (BioProject PRJNA870170)

https://www.ncbi.nlm.nih.gov/bioproject/?term=prjna870170

## Author statements

### Author contributions

L.S.K. – conceptualization, investigation, data curation, analysis and visualisation, writing original draft. M.S. – conceptualization, investigation, data analysis, writing- review and editing. M.F. – bioinformatic analysis (metagenomic sequencing), data curation and analysis, writing- review and editing. L.M.J.- metagenomic sequencing, writing- review and editing. J.A.L. - bioinformatic analysis, writing- review and editing. J.S. – resources, writing- review and editing. S.L.B. – methodology, writing- review and editing. B.P.H. – conceptualization, writing- review and editing, funding. N.L.S. – conceptualization, writing- review and editing, funding.

### Ethical approval

Ethical approval for this study was obtained from the University of Melbourne Human Research Ethics Committee under the project “Pathogen Recovery in Human Clinical Samples” (Reference Number: 2021-22158-20892-3).

## Acknowledgements

We acknowledge and thank the diagnostic laboratories in the state of Victoria for referring clinical specimens that were PCR-positive for *Bordetella* spp. to Microbiological Diagnostic Unit Public Health Laboratory (MDU PHL) for this study. The authors gratefully acknowledge all MDU PHL staff who involved in processing, characterisation and sequencing of the samples used in this study. The contributing laboratories included Australian Clinical Labs, Dorevitch Pathology, Monash Health Pathology, Austin Pathology, Melbourne Pathology and The Royal Melbourne Hospital.

