## Supplementary Methods for "Enhancing molecular and genomic approaches for *Bordetella pertussis* investigation and public health surveillance in Victoria, Australia"

Supplementary Method 1 – Real-time PCR for *Bordetella* spp. detection and differentiation

Supplementary Method 2 – Multiplex PCR amplicon sequencing for *Bordetella pertussis* lineage typing

### Supplementary Method 1 – Real-time PCR for *Bordetella* spp. detection and differentiation

The multiplex real-time PCR assay for *Bordetella* spp. detection and differentiation was performed as previously described [10]. Primer and probe sequences for all targets are provided in Table SM1.1.

**Table SM1.1:** **Sequences of primers and probes utilised for real-time PCR**

| **Target** | **Primer/Probe** | **Sequence (5’-3’)** |
| --- | --- | --- |
| *IS481* | Forward | GCCGGATGAACACCCATAAG |
|  | Reverse | GCGATCAATTGCTGGACCAT |
|  | Probe | 6FAM – CGATTGACCTTCCTACGTC – MGBNFQ |
| *IS1001* | Forward | AATTGCTGCAAGCCAACCA |
|  | Reverse | CCAGAGCCGTTTGAGTTCGT |
|  | Probe | VIC – ACATAGACCGTCAGCAG – MGBNFQ |
| *IS1002* | Forward | CTAGGTCGAGCCCTTCTTGTTAAC |
|  | Reverse | GCGGGCAAGCCACTTGTA |
|  | Probe | VIC – CATCGTCCAGTTCTGTTGCATCACCC – QSY |
| *recA* | Forward | CGGTTCGCTGGGTCTCG |
|  | Reverse | CCCGCGGCAGACCAC |
|  | Probe | 6FAM – CATCGCATTGGGCG – MGBNFQ |

Two multiplex real-time PCR assays were performed: one targeting IS*481* and IS*1001*, and a second targeting *recA* and IS*1002*. Each 50 μL reaction contained 0.3 μM of each primer, 0.2 μM of each probe, 1× SensiFAST™ Probe Lo-ROX (Bioline), 1× qPCR Extraction Control RED – Control Mix 25× (Meridian Bioscience) and 5 μL of extracted DNA. Amplification was performed on the QuantStudio™ 5 Real-Time PCR System (Thermo Fisher Scientific) with an initial denaturation at 95°C for 3 min, followed by 45 cycles of 15 seconds at 95°C and 1 min at 60°C. Cycle threshold (Ct) values were determined automatically using the QuantStudio™ Design and Analysis Software v1.5.3 (Thermo Fisher Scientific).

*Bordetella* spp. identification criteria based on the detection profile of IS*481*, IS*1001*, IS*1002*, and *recA* are provided in Table SM1.2.

| **PCR target** | | | | **Species assignment** |
| --- | --- | --- | --- | --- |
| **IS*481*** | **IS*1001*** | **IS*1002*** | ***recA*** |  |
| Detected | Not detected | Detected | Not detected | *B. pertussis* |
| Not detected | Detected | Detected | Not detected | *B. parapertussis* |
| Detected | Not detected | Not detected | Detected | *B. holmesii* |
| Detected | Not detected | Not detected | Not detected | *Bordetella* spp. |
| Not detected | Detected | Not detected | Not detected | *Bordetella* spp. |

**Table SM1.2: PCR target profiles and species assignment criteria**

### Supplementary Method 2 – Multiplex PCR amplicon sequencing for *Bordetella pertussis* lineage typing

Multiplex PCR amplicon sequencing was performed using previously described primer sets targeting lineage-associated loci [12]. Primer sequences were identical to those reported previously and are provided in Table SM2.

The first-round multiplex PCR was performed in a 20 µL reaction containing 4 µL of 5 M betaine, 2 µL of 10× DreamTaq Buffer, 0.4 µL of 10 mM dNTP mix, 0.7 µL each of forward and reverse primer mix, 0.2 µL of 5 U/µL DreamTaq DNA Polymerase (Thermo Scientific), 2 µL of water and 10 µL of extracted DNA. Primer concentrations in the forward and reverse primer mixes and the final multiplex PCR are provided in Table SM2. Multiplex PCR cycling conditions were an initial denaturation at 95°C for 1 min, followed by 30 cycles of denaturation at 95°C for 30 s, primer annealing at 60°C for 30 s, and extension at 72°C for 1 min, with a final extension at 72°C for 7 min.

Following first-round multiplex PCR, amplicons were purified using 0.8× AMPure XP beads (Beckman Coulter). A second-round PCR was performed in a 50 µL reaction containing 25 µL of 2× KAPA HiFi HotStart ReadyMix (Roche), 10 µL of IDT for Illumina Unique Dual Indexes (Plate A/Set 1 Index Adapters - <https://support-docs.illumina.com/SHARE/AdapterSequences/Content/IDTIllumina-UDIndexes.htm>) and 15 µL of purified PCR product from the first-round PCR. Indexed libraries were purified using 0.8× AMPure XP beads, pooled, and sequenced on an Illumina iSeq 100 with iSeq 100 i1 Reagent v2 (300-cycle) to generate 2 × 150 bp paired-end reads.

| **Target** | **Primer** | **Primer sequence (5' --> 3')*** | **Concentration in primer mix (µM)** | **Final concentration in multiplex PCR (µM)** |
| --- | --- | --- | --- | --- |
| *ptxP* | ptxP_F | TCGTCGGCAGCGTCAGATGTGTATAAGAGACAGCTTCCGTACATCCCGCTACT | 7.0 | 0.25 |
|  | ptxP_R | GTCTCGTGGGCTCGGAGATGTGTATAAGAGACAGGCGTTTTGATGGTGCCTATT | 7.0 | 0.25 |
| *fim3* | fim3_F | TCGTCGGCAGCGTCAGATGTGTATAAGAGACAGAACTGCCCAAGATTTCCAAG | 9.9 | 0.35 |
|  | fim3_R | GTCTCGTGGGCTCGGAGATGTGTATAAGAGACAGGGATGCGTTGTAGGTCTGCT | 9.9 | 0.35 |
| *fhaL* | fhaL_F | TCGTCGGCAGCGTCAGATGTGTATAAGAGACAGGAACAGCACGACCTCCAACT | 9.9 | 0.35 |
|  | fhaL_R | GTCTCGTGGGCTCGGAGATGTGTATAAGAGACAGGCGAGATGGCATTCAGGT | 9.9 | 0.35 |
| *gor* | gor_F | TCGTCGGCAGCGTCAGATGTGTATAAGAGACAGATCGAGGTGGACGACGAATA | 9.9 | 0.35 |
|  | gor_R | GTCTCGTGGGCTCGGAGATGTGTATAAGAGACAGTCTGGGTCAGGGTCAGTTTC | 9.9 | 0.35 |
| *livJ* | livJ_F | TCGTCGGCAGCGTCAGATGTGTATAAGAGACAGGACGATCTCGGCGTTGAC | 12.7 | 0.44 |
|  | livJ_R | GTCTCGTGGGCTCGGAGATGTGTATAAGAGACAGGTACTACGGCGGCTATCACC | 12.7 | 0.44 |
| *dapB* | dapB_F | TCGTCGGCAGCGTCAGATGTGTATAAGAGACAGCGCCAAACAGGAAGAAAAAG | 15.5 | 0.54 |
|  | dapB_R | GTCTCGTGGGCTCGGAGATGTGTATAAGAGACAGACGGCTTCGATCAGCATCT | 15.5 | 0.54 |
| *pykA* | pykA_F | TCGTCGGCAGCGTCAGATGTGTATAAGAGACAGGCTGAACAAGGTGGTCATCA | 18.3 | 0.64 |
|  | pykA_R | GTCTCGTGGGCTCGGAGATGTGTATAAGAGACAGGTGGTGGTAGGGTGCTTCTC | 18.3 | 0.64 |
| *BP2072* | BP2072_F | TCGTCGGCAGCGTCAGATGTGTATAAGAGACAGGACCTGCTGGTCAACTTCAA | 9.9 | 0.35 |
|  | BP2072_R | GTCTCGTGGGCTCGGAGATGTGTATAAGAGACAGCGCCTTGTCGAGATCGTT | 9.9 | 0.35 |
| *fhaB* | fhaB_F | TCGTCGGCAGCGTCAGATGTGTATAAGAGACAGGGGGAGCTGACCGTCAAGGC | 7.0 | 0.25 |
|  | fhaB_R | GTCTCGTGGGCTCGGAGATGTGTATAAGAGACAGATCCAGGTGCATGTCCTCGC | 7.0 | 0.25 |

**Supplementary Method Table SM2: Primer sequences used for multiplex PCR amplicon sequencing**

*Red and green sequences indicate Illumina Nextera adapter overhangs added to target-specific primers for subsequent indexing during the second-round PCR using IDT for Illumina Unique Dual Indexes.
